# Hypertension and Diabetes Delay the Viral Clearance in COVID-19 Patients

**DOI:** 10.1101/2020.03.22.20040774

**Authors:** Xiaoping Chen, Wenjia Hu, Jiaxin Ling, Pingzheng Mo, Yongxi Zhang, Qunqun Jiang, Zhiyong Ma, Qian Cao, Liping Deng, Shihui Song, Ruiying Zheng, Shicheng Gao, Hengning Ke, Xien Gui, Åke Lundkvist, Jinlin Li, Johanna F Lindahl, Yong Xiong

## Abstract

**Objectives:** Comorbidities have significant indications for the disease outcome of COVID-19, however which underlying diseases that contribute the most to aggravate the conditions of COVID-19 patients is still largely unknown. SARS-CoV-2 viral clearance is a golden standard for defining the recovery of COVID-19 infections. To dissect the underlying diseases that could impact on viral clearance, we enrolled 106 COVID-19 patients who were hospitalized in the Zhongnan Hospital of Wuhan University, Wuhan, China between Jan 5 and Feb 25, 2020.

**Methodology:** We comprehensively analyzed demographic, clinical and laboratory data, as well as patient treatment records. Survival analyses with Kaplan-Meier and Cox regression modelling were employed to identify factors influencing the viral clearance negatively.

**Results:** We found that increasing age, male gender, and angiotensin-converting enzyme 2 (ACE2) associated factors (including hypertension, diabetes, and cardiovascular diseases) adversely affected the viral clearance. Furthermore, analysis by a random forest survival model pointed out hypertension, cortisone treatment, gender, and age as the four most important variables.

**Conclusions:** We conclude that patients at old age, males, and/or having diseases associated with high expression of ACE2 will have worse prognosis during a COVID-19 infections.

## Introduction

Emerging infectious diseases (EID) in both animals and humans cause major economic and health burden globally and around 75% of all EIDs are zoonotic, of which many originate from wildlife [1]. There has been an estimate that a new human disease appears every four months, driven by mainly anthropogenic changes, including land-use change (deforestation, irrigation, urbanization), climate change, population growth, and globalization [1, 2]. The most noteworthy and important EIDs during this millennium belong to the genus coronavirus (CoV), a group of enveloped, single-stranded, positive-sense RNA viruses. In 2002-03, an outbreak of severe acute respiratory syndrome (SARS), caused by a coronavirus later termed SARS-CoV, started out of China. This was followed by the spread of Middle East respiratory syndrome coronavirus (MERS-CoV) in 2012. In 2019, a new pandemic started out of China, COVID-19, caused by SARS-CoV-2 [3-5]. The origins of these zoonotic viruses seem to be bats, which have been shown to harbor many different coronaviruses [6-8], even though it seems necessary with an amplifying host closer to humans to obtain a species jump to man. For MERS-CoV, the main amplifying host close to humans has been identified as dromedaries, while for SARS-CoV-2, the amplifiers were likely small mammals sold at live animal markets [3–5]. Cross-species transmission and the ability to sustain many cycles of human-to human transmission requires a solid interaction between the receptor-binding viral proteins and receptors on the host cells [9]. The host cell receptor for SARS-CoV and SARS-CoV-2 seems to be the angiotensin-converting enzyme 2 (ACE2) enzyme, presented on many epithelial cells in the respiratory tract [10-12].

COVID-19 most often displays as fever, cough and mild fatigue, sometimes also dyspnea, myalgia and severe anorexia, and may develop into critical respiratory failure that sometimes becomes fatal [13-16]. In one study, mild cases seemed to account for 81%, while another study found at least 50% of the infected were asymptomatic, but transmission can occur from both symptomatic and asymptomatic carriers [15, 17, 18]. As compared to SARS-CoV with a reproductive number (R0) of 2-5 [19, 20] and R0 of MERS-CoV from 0.6 to 3 [21], it seems that SARS-CoV-2 is likely having an R0 in the range of 2-3 [20], but the new virus has already killed more people due to its global spread [22].

According to the clinical characteristics, It has been shown that severe COVID-19 often occurred in patients with diabetes, hypertension or other comorbidities [11]. In COVID-19 patients with underlying diseases, a very high case fatality rate (73.3%) has been observed [23]. However, how these comorbidities affect the COVID-19 prognosis is not yet understood. Potential factors might be that these comorbidities directly accelerate the damage of target tissues or that they favour the virus life-cycle during a SARS-CoV-2 infection. Trying to answer this critical and complex question, we analyzed data from COVID-19 hospitalized patients and evaluated which underlying diseases that had the highest risk to aggravate the conditions of a COVID-19 infection. We found that chronic diseases including diabetes and hypertension deteriorate the clearance of SARS-CoV-2.

## Methods

Between Jan 5 and Feb 25, 2020, we collected the records of 106 hospitalized patients, whose throat-swab specimens had been tested positive for SARS-CoV-2 by qRT-PCR on admission according to a protocol previously described [13]. The patients who repeatedly tested SARS-CoV-2 RNA negative for at least two times with an interval of at least one day were regarded as viral negative. The admission date was used as the starting time-point for the viral clearance process, and the date of the second negative detection of viral RNA was calculated as the end time-point of viral clearance. We recorded patient demographic data including gender, age and lag time from onset of disease until seeking hospital care (lag time); clinical data of chronic diseases including hypertension, chronic obstructive pulmonary disease (COPD), gender, age, liver disease, cancer, cardiovascular diseases and diabetes existed as comorbidities; treatment data including usage of antivirals or cortisone. The data were imported and analyzed using R language, implemented in RStudio [24, 25]. Package survival, survminer, ranger and ggplot2 were employed for survival analysis, modelling and visualization separately [26-30]. Informed consents were obtained from all patients upon admission to the Department of Infectious Diseases, Zhongnan Hospital of Wuhan University, Wuhan, China. This study was approved by the ethics board in Zhongnan Hospital of Wuhan University, Wuhan, China (No.2020011).

## Results

In a total of 106 patients, 63 females (63/106, 59.4%) and 43 males (43/106, 40.6%), were included in the study. The days of observation spanned from 4 days to 37 days. On day 37, 99 patients had cleared the virus infection and 5 had died (4.7%, 95% confidence interval 1.5-10.7%). The age of the patients spanned from 20 to 96 years with an average of 50.9 years (25%–75% interquartile range (IQR), 35.0 – 64.6 years). Underlying diseases included 17 patients with hypertension (16.0%), six with cardiovascular disease (5.7%), nine with diabetes (8.5%), two with malignancy (1.9%) and 10 with chronic liver disease (9.4%). Antivirals and cortisone had been used in 67 (63.2%) and 54 (50.9%) of the patients, respectively.

We monitored the time until viral clearance during the clinical course of the patients. The patients had a median time of 15 days (IQR, 10 – 20 days) until negative of SARS-CoV-2 viral RNA. The overall viral clearance events by using Kaplan-Meier survival analysis are summarized in Figure. Over 80% patients reached viral clearance (qRT-PCR was negative for SARS-CoV-2) within 25 days from the first day of laboratory-confirmed SARS-CoV-2 infection.

In another Cox proportional hazards model, we included the demographic, clinical, and treatment variables to model the risk factors for viral positivity over time. We found hypertension (P=0.01) and corticosteroid treatment (P=0.03) to be risk factors preventing viral clearance. To better observe the contribution of each variable to the Cox model over time, Aalen’s additive regression model was used to plot the additive coefficient of each variable over the time. Within the first five days of the viral clearance process, no variable had any effect on the model (coefficient fluctuates around 0). With longer time of observation (from day 5 to day 20), COPD, age, carcinoma, corticosteroid treatment, diabetes, gender, hypertension, and lag time had negative effects (coefficient < 0) on the model, suggesting that if patients had those factors, the time of viral clearance were prolonged. However, the interval of additive coefficient increased over time, suggesting that an uncertainty of each variable’s contribution to the model is increased due to a reduced number of patients remaining in the model.

Finally, a random forest survival model was used to analyze which variables explained SARS-CoV-2 viral positivity. We found that hypertension (index=0.0353), the usage of cortisone (index=0.0265), gender (index=0.0221), and age (index=0.0076) were the top four important variables. By examining the vibration of the risks function over time, three survival models: Kaplan-Meier, Aalen’s additive regression model and random forest survival model were used to model when the patients become clear from the viral infections. Interestingly, we found hypertension (in three models), treatment with cortisone (in three models), increasing age (in the random forest survival model), and male gender (in the random forest survival model) had negative effects on the viral clearance in COVID-19 patients.

## Discussion

Our analyses pinpointed hypertension as the most important risk factor and this has been suggested by other studies as well [11, 23]. Hypertension, diabetes, and cardiovascular diseases are associated with abnormal regulation of renin-angiotensin system (RAS) and ACE2 has a key role involved in this process [11]. It is likely that the patients with these comorbidities had been treated with ACE2 inhibitors, which could have affected the presence of these receptors, increasing the receptor usage of SARS-CoV-2 [10]. Other studies have showed that high expression of ACE2 in the patients with hypertension, diabetes, and cardiovascular diseases might facilitate SARS-CoV-2 to enter the targeted cells in the respiratory system, and prolong the time of viral clearance [11, 31]. On the other hand, old people usually have a high risk of in hypertension, diabetes, and cardiovascular diseases [32], making age and these diseases difficult to discern.

Sex, age and immunity are notable biological factors during combating infectious pathogens [33]. In our model, male and old age were also identified as risk factors. The immune system is different between females and males [34]. Females may have lower susceptibility to viral infections, because estrogen and progesterone can help to increase the innate and adaptive immune responses, and many immune genes are X-linked [35]. High immune reactivity post viral infection in women can accelerate the process of viral clearance but, on the other hand, could be leading to immune-pathogenicity and autoimmunity. Gender differences can also affect risk factors, with men e.g. being more likely to develop hypertension [36]. Males have a high ratio of hypertension in mice models due to decreased adipose ACE2 activity [37]. However, although the ACE2 gene is located in the X chromosome, there is no evidence of different expression of ACE2 between the sexes [38].

Until now, there is no effective medicine available to treat COVID-19. Accordingly, the usage of antivirals (Arbidol) had no effect on the viral clearance, according to our results. The usage of cortisone had unexpectedly a negative effect on the process of viral clearance, although cortisone was commonly used in the SARS patients [39]. However, it is possible that cortisone was targeted at particularly severe cases, which may have confounded the results. Compared to SARS, the outcome of using corticosteroids in COVID-19 patients is still unclear [14, 40]. Given that corticosteroids as immune-modulators that can decline circulating specific B- and T-cell subsets [41] and our modelling, the usage of cortisone would not be recommended.

In conclusion, patients with hypertension, male and old age have increased risk for late viral clearance of the virus, and thus worsened prognosis in COVID-19 infections, likely due to the fact that those risk factors are associated with high expression of ACE2. The usage of ACE2 inhibitors for patients with hypertension, diabetes, and CVD need to be carefully considered.

It is recommended that future projects not only collect data on comorbidities, but also compare the risk for patients with the same disease in regards to the medications they were taking before the infection by SARS-CoV-2. The general profile of hazards in COVID-19 hospitalized patients over time could provide us more information in the decision-making and personalized treatment during the clinical practice.

## Data Availability

All data referred to in the manuscript are availability.

## Acknowledgements

This study was funded by the Zhongnan Hospital of Wuhan University, Science, Technology and Innovation Seed Fund, grant number znpy2018007 and the Swedish Research Council, grant number 2017-05807. The funders had no role in study design, data collection or analysis, decision to publish or preparation of the manuscript. The authors declared no competing interests.

---

**Figure 1.**
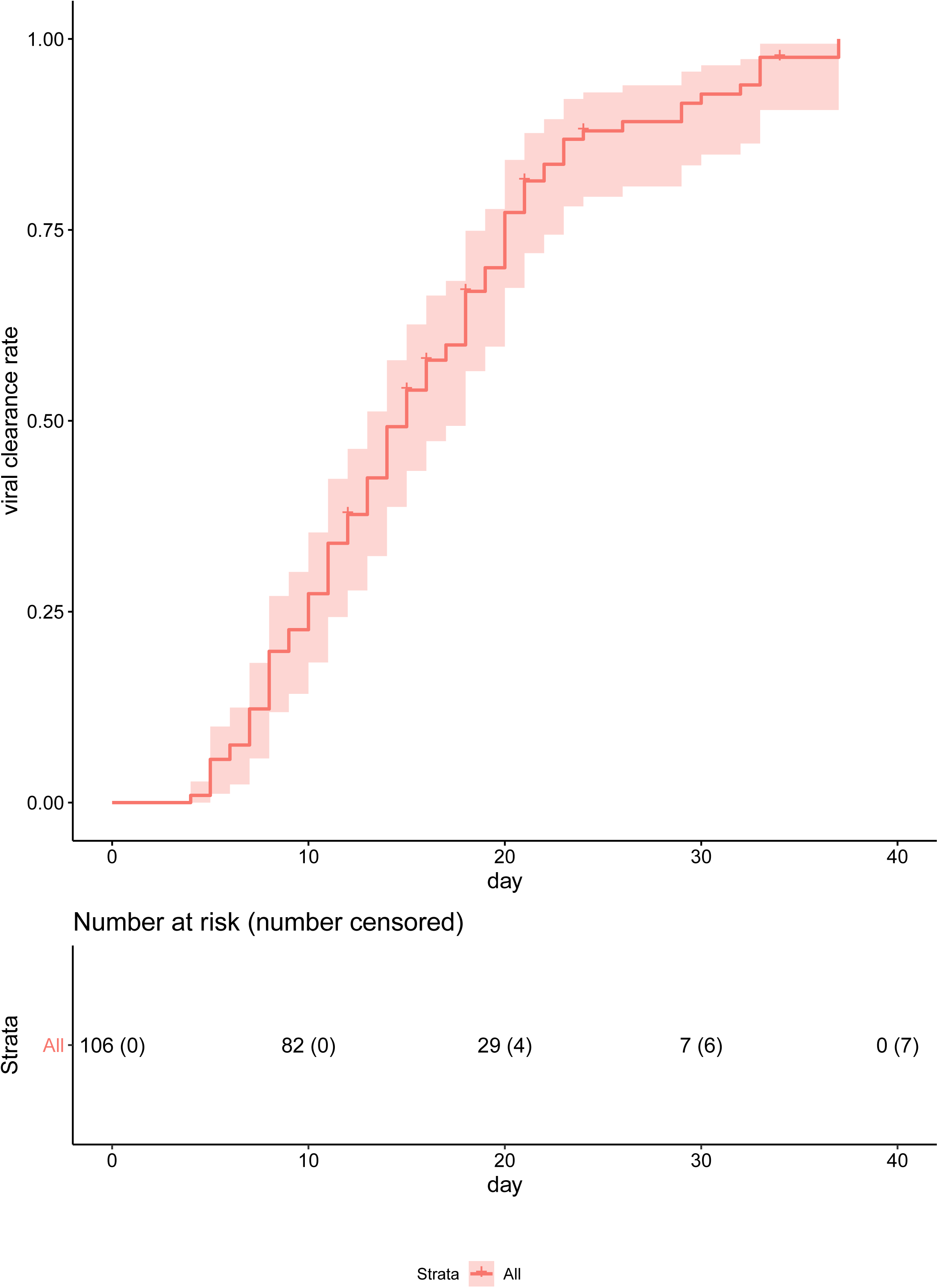
Kaplan-Meier estimates of SARS-CoV-2 viral clearance rate over time.

**Figure 2.**
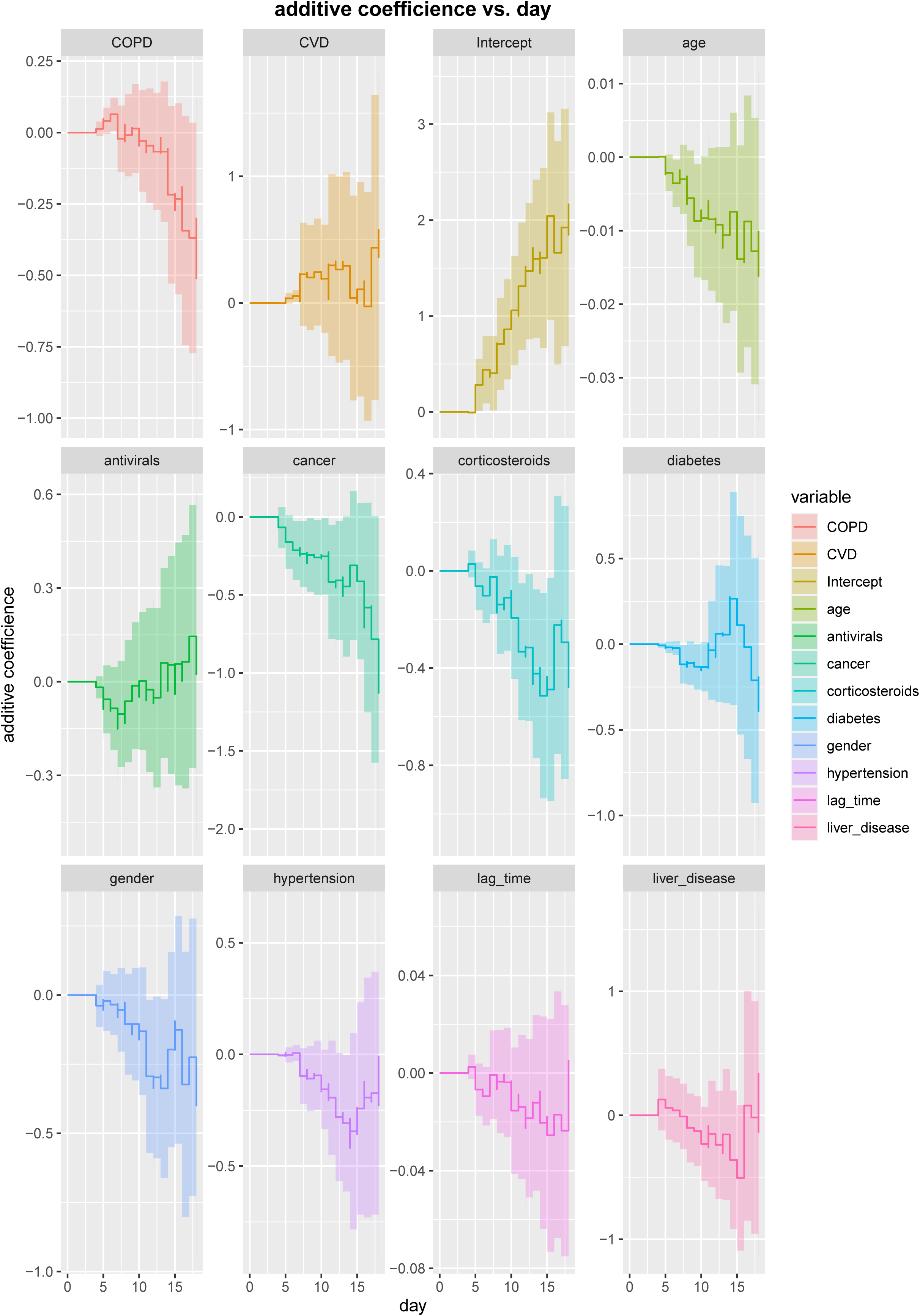
Aalen’s additive regression model of each covariant over time. Covariants included gender, age, lag time, hypertension, COPD, liver diseases, diabetes, CVD, cancer, antivirals and corticosteroids as treatment.

